# Comparison study of Olmesartan and Valsartan On myocardial metabolism In patients with Dilated cardiomyopathy (OVOID) trial

**DOI:** 10.1101/2024.12.12.24318958

**Authors:** Sangyong Jo, Kyungil Park, Jae Hyuk Choi, Chang-Bae Sohn, Jeonghwan Kim, Yong-Seop Kwon, Su Hong Kim, Tae-ho Park

## Abstract

**Background:** Myocardial metabolism plays an important role in maintaining cardiac function. Patients with dilated cardiomyopathy (DCMP) exhibit alterations in myocardial metabolism characterized by increased myocardial glucose metabolism. This study aimed to evaluate effects on myocardial metabolism of angiotensin-II receptor blockers, olmesartan and valsartan, in patients with DCMP.

**Methods:** OVOID (a comparison study of Olmesartan and Valsartan effects on myocardial metabolism in patients with Dilated cardiomyopathy), an investigator-initiated, multicenter, randomized controlled study of DCMP patients with New York Heart Association (NYHA) class II-IV, was conducted. The primary outcome was myocardial glucose metabolism measured by standardized uptake value ratio (SUVR) at 6 months after treatment. To measure SUVR, ^18^F-fluoro-2-deoxyglucose (FDG) cardiac positron emission tomography (PET) was performed at baseline and six months after receiving the study agent. A total of 44 patients were randomized at a 1:1 ratio to receive olmesartan (at a dose of 20 mg once daily) or valsartan (at a dose of 160 mg twice daily) for 6 months, in addition to a recommended therapy.

**Results:** Baseline clinical characteristics and SUVR measured by ^18^F-FDG PET data did not differ significantly between olmesartan and valsartan groups. The average left ventricular ejection fraction (LVEF) of patients was 25.1±7.8. Among patients with DCMP who received olmesartan or valsartan for 6 months, LVEF significantly increased. However, it was not significantly different between the two groups. Six-month follow-up ^18^F-FDG PET showed that SUVR value was significantly lower in the olmesartan group than that in the valsartan (3.76 ± 2.12 versus 7.33 ± 4.08, *P* = 0.01).

**Conclusions:** Six months of olmesartan therapy significantly decreased myocardial glucose metabolism in DCMP patients compared to valsartan therapy for six months.

**Trial registration:** ClinicalTrials.gov; NCT04174456; 18 November 2019

**Clinical Perspective:** **What is New?**

- In patients with dilated cardiomyopathy (DCMP), six months of olmesartan therapy significantly decreased myocardial glucose metabolism in DCMP patients compared to valsartan therapy for six months.
- Olmesartan is as effective as valsartan in treating heart failure (HF) and improving left ventricular ejection fraction and symptoms in DCMP.

**What Are the Clinical Implications?**

Myocardial metabolism plays an important role in maintaining cardiac function. Impairment of myocardial metabolism can contribute to progression of left ventricular remodeling and contractile dysfunction in HF. Results of the present study revealed different effects on myocardial glucose metabolism between angiotensin II receptor blockers (ARBs) in DCMP. In addition to its inhibitory effects on renin-angiotensin-aldosterone systems in HF, olmesartan as an ARB might be considered when considering its effects on myocardial glucose metabolism. These findings provide further insights into mechanisms responsible for the beneficial effect of ARB on myocardial metabolism. They could redirect future translational investigations in an effort to identify novel therapeutic targets for myocardial recovery in patients with DCMP.

## Introduction

Myocardial metabolism plays an important role in maintaining cardiac function.^1,2^ Metabolic changes in a failing heart are linked to functional and structural changes.^3,4^ Patients with dilated cardiomyopathy (DCMP) may exhibit alterations in myocardial metabolism that can lead to progressive impairment of cardiac high-energy phosphate production with a metabolically inefficient heart.^5–7^ Recent evidences suggest that angiotensin II might be involved in these perturbations in cardiac energy metabolism.^8,9^ Overactivation of the renin-angiotensin-aldosterone system (RAAS) is one of the key detrimental mechanisms of DCMP progression. It is associated with poor prognosis. As a result, suppressing the RAAS might positively affect myocardial metabolism in DCMP patients.^10,11^ Thus, suppression of RASS is a potential therapeutic strategy in patients with DCMP. Although angiotensin II receptor blockers (ARBs) have been shown to be effective in inhibiting the RAAS, there has been no direct comparison of differential effects of these agents on myocardial metabolism.^12,13^ Thus, the purpose of this study was to compare effects of olmesartan and valsartan on myocardial glucose metabolism in DCMP patients. In this study, we used cardiac positron emission tomography (PET) to evaluate myocardial glucose metabolism in patients with DCMP.

## Methods

Data that support findings of this study are available from the corresponding author upon reasonable request.

### Study population

Patients were eligible for this study if they were diagnosed with DCMP with a left ventricular (LV) ejection fraction (EF) < 40% and an LV end-diastolic diameter > 117% of the predicted value corrected to body surface area and age with New York Heart Association (NYHA) functional classes of II and IV. A diagnosis of DCMP was determined using the currently accepted criteria.^14^

Exclusion criteria were: 1) less than 20 years or more than 85 years old, 2) the presence of hemodynamic instability, 3) known intolerance to olmesartan and valsartan, 4) coronary artery disease based on coronary angiography (CAG) (≥ 50% stenosis in ≥ 1 of the major coronary arteries) and/or a history of myocardial infarction or angina pectoris, 5) acute or subacute stage of myocarditis, 6) primary valve disease, 7) excessive use of alcohol, 8) expected or performed cardiac resynchronization therapy, and heart transplantation, 9) stress-provoked Takotsubo cardiomyopathy, 10) tachycardia-induced cardiomyopathy, 11) peripartum cardiomyopathy, 12) Cor pulmonale, 13) impaired renal function (estimated glomerular filtration rate < 60 ml/min/1.73m^2^, 14) a life expectancy of less than one year, or 15) inability to provide informed consent.

### Study endpoints

The primary outcome was myocardial glucose metabolism as measured by standardized uptake value ratio (SUVR) at six months after treatment. Secondary endpoints included changes in SUVR, N-terminal pro B-type natriuretic peptide (NT-proBNP) levels, LVEF, and occurrence of predefined clinical events after receiving the study agent. In this study, clinical events were all causes of death, cardiovascular death, left ventricular assist device implantation, listing for cardiac transplantation, hospitalization for worsening heart failure (HF), and intensification of therapy defined by an increase in the diuretic dose > 50% in an outpatient setting during six months of follow-up.

### Trial design and oversight

OVOID (a comparison study of Olmesartan and Valsartan effects on myocardial metabolism in patients with Dilated cardiomyopathy) was a prospective, multicenter, randomized, open-label, active-controlled study that enrolled patients with first diagnosed with DCMP within the last 4 weeks for NYHA class II-IV. This study was conducted across seven sites in South Korea. Participating sites and investigators are listed in Appendix S2 of online-only Data Supplement. The recruitment of participants started from December 2019 and ended in December 2021. Study approval was granted by the Ethics Committee/Institutional Review Board of Dong-A University Hospital. All patients provided written informed consent. The trial was overseen by an independent data monitoring committee.

Before administration of the first dose of olmesartan or valsartan, patients underwent a physical exam, trial-related laboratory assessments, and coronary angiography. ^18^F-fluorodeoxyglucose (^18^F-FDG) PET was used for measuring SUVR as a baseline.^15^ Patients confirmed eligible were randomized at 1:1 to olmesartan 20 mg once daily or valsartan 160 mg twice daily (initially at a dose of 40 mg twice daily, which was increased to 160 mg twice daily as tolerated throughout the study period). In addition, the guideline directed standard of care therapy for six months except for angiotensin converting-enzyme (ACE) inhibitor and ARB.^16,17^ At week 24, ^18^F-FDG PET and echocardiography were performed by a core laboratory staff blinded to random assignment. The dose of the study drug could be reduced for patients who were not clinically feasible at target dose. Patients then entered a 6-month treatment period, during which they were followed via 5 in-person study visits (at 2, 4, 8, 16 and 24 weeks), at which time physical exams and laboratory assessments were repeated and assessment of drug compliance was completed. Patient’s NYHA functional class and predefined clinical events were recorded at each clinical visit. Adverse events were reported from in-hospital observation and follow-ups. At each visit, patients were interviewed about the occurrence of any adverse events, including time of onset and duration. The study population was instructed to contact the research staff via mobile phone at any time if adverse events occurred during the study period.

### ^18^F-FDG PET measurements

^18^F-FDG PET imaging was performed at the core lab (Dong-A University Hospital, Busan, South Korea). All studies were performed with a Biograph mCT flow scanner (Siemens Healthcare, Knoxville, TN, USA) consisting of a PET scanner coupled with a multi-detector CT scanner, allowing acquisition of co-registered CT and PET images at the same time. After fasting for at least six hours, different doses of insulin (1U-5U) were injected after 40 – 60 minutes of oral administration of 25 – 50 g of a 50% glucose load according to different blood glucose levels for patients to reach an optimal blood glucose concentration of 5.55 – 7.77 mmol/L.^18^ After confirming that the proper blood sugar level was reached, low-dose CT was acquired, first with 100 kV, 30 mAs, 0.5 s each rotation, and 3.75 mm of slice thickness, without oral or intravenous contrast media before PET acquisition. Simultaneously with the initiation of PET acquisition, 185 MBq of ^18^F-FDG was rapidly injected intravenously. PET scan was acquired for 50 min in a list-mode. Static, ECG-gated, and dynamic PET images were generated from list-mode PET data. The first two PET images (static and ECG-gated images) were reconstructed using the last 10 min of the entire data acquisition. Dynamic PET images were also reconstructed in 12 frames of 10 seconds, 3 frames of 20 seconds, 6 frames of 40 seconds, 4 frames of 60 seconds, and 8 frames of 5 minutes, with a total of 33 frames. These frames were generated with a 200 × 200 matrix, 2-mm slice thickness, a Gaussian filter of 4 mm in full width at half maximum, three iterations, and 21 subsets.

Dynamic PET data were analyzed using PMOD software (version 3.6, PMOD Technologies LLC., Zurich, Switzerland) to measure SUVR as myocardial metabolism assessment.^19^ The PET image was automatically loaded using the PMOD Cardiac PET modeling module to align directions. A total of 18 volumes of interest (VOIs) including 17 segments of the LV were manually oriented and adjusted. They were then automatically segmented using the PCARD model in PMOD to measure standardized uptake value (SUV). The SUV was calculated from the activity concentration (kBq/mL) divided by total administered activity (kBq) within the phantom background chamber and normalized to the weight (g) of the solution in the phantom.^20^ In this study, the SUVR was defined as the SUV of the myocardial VOI divided by the SUV of the left atrial blood pool VOI. These analyses were carried out by two nuclear medicine specialists with experience in ^18^F-FDG PET who were blinded to treatment assignment.

### Statistical analysis

The primary hypothesis to be tested was differential effect of olmesartan on myocardial glucose metabolism in DCMP patients compared to valsartan with a six-month follow-up period. We calculated the number of study subjects based on studies using myocardial glucose metabolism in DCMP patients.^6,21–24^ The sample size was calculated using the study’s primary objective to confirm a 20% relative difference in myocardial glucose metabolism between olmesartan and valsartan treatments with a power of 90%. We expected that the relative difference in myocardial glucose metabolism would be at least 20% in DCMP patients. For the primary end point, a sample size of 20 for each group was projected to achieve 90% power with α = 5% to detect a 20% relative difference in SUVR between the two groups. With an expected drop-out rate of approximately 10%, the final sample size was estimated to be 22 patients per treatment group.

Statistical analyses were performed on an intention-to-treat basis. Continuous variables are presented as mean ± SD or median and interquartile range (IQR). They were compared by Student’s t-test or Mann-Whitney U test, respectively, depending on the normality of variables. Chi-squared test was used to analyze categorical variables. Repeated measures between baseline and follow-up were evaluated using paired t-test for continuous Gaussian-distributed parameters or the Wilcoxon test, as appropriate. Spearman’s correlation analysis was used to analyze correlations between SUVR and NT-proBNP or LVEF. The association between SUVR and possible influencing factors was also examined with a linear regression model. Independent influencing factors of SUVR were analyzed by multivariate logistic regression. Safety analysis included calculation of frequencies and rates of adverse events reported in the two groups. All statistical analyses were performed using SPSS Version 16.0 or higher. Statistical significance was considered when *P*-value was less than 0.05.

## Results

### Study patients

From December 2019 to December 2021, a total of 263 patients were screened, of which 44 patents from 7 sites were enrolled. A total of 44 patients were included in the intention-to-treat analyses. New York Heart Association (NYHA) class II symptoms were present in 20.5%, with class III/IV symptoms present in 79.5%. Characteristics of patients in the two study groups were similar at baseline (Table 1). The olmesartan group and the valsartan group showed no significant difference in NT-proBNP (2295, IQR 1050–5461 versus 2720, IQR 1084–6044 pg/mL, *P* = 0.42). All patients received recommended pharmacologic therapy for heart failure (HF) with reduced EF. Medication uses for HF were not different between the two groups (Table 1). Figure 1 shows random assignment and follow-up of patients. One patient in each group died during the study. Premature permanent treatment discontinuation occurred in one patient in the olmesartan group and 2 patients in the valsartan group, resulting in 20 patients in each group completing the trial of treatment. Follow-up medication use was similar in the two groups at 6 months. In the valsartan group, twenty (90.9%) patients took the maximum dose (320 mg/d) of valsartan. Both systolic and diastolic blood pressures (BP) decreased substantially by almost the same extent in both olmesartan and valsartan groups, showing no inter-group difference during the treatment (systolic/diastolic BP at baseline and 6 months: 128.5 ± 17.8/76.4 ± 12.9 mmHg and 112.8 ± 13.7/74.2 ± 10.1 mmHg, respectively, in the olmesartan group, and 126.3 ± 16.4/77.8 ± 11.5 mmHg and 110.1 ± 12.2/73.6 ± 9.8 mmHg in the valsartan group). Safety follow-up was competed in all patients. No patient withdrew consent or was lost to follow-up.

**Figure 1.**
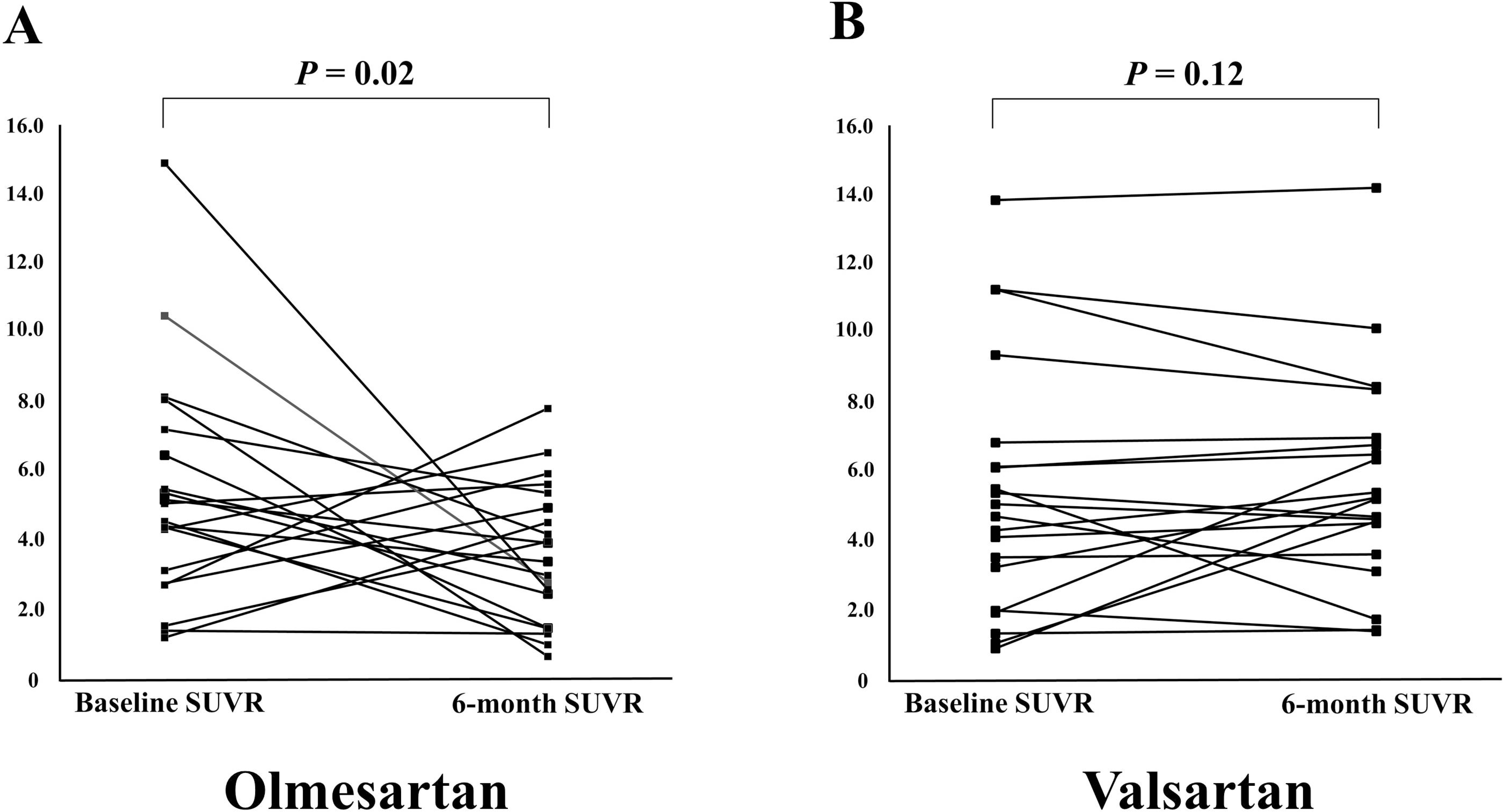
Randomization and follow-up. The intention-to-treat population included all the patients who had undergone randomization with valid informed consent and who had received at least one dose of study medication. Of the 44 patients who ultimately underwent randomization, premature permanent treatment discontinuation occurred in one patient in the olmesartan group and 2 patients in the valsartan group, resulting in 20 patients in each group completing the trial of treatment.

**Table 1.**
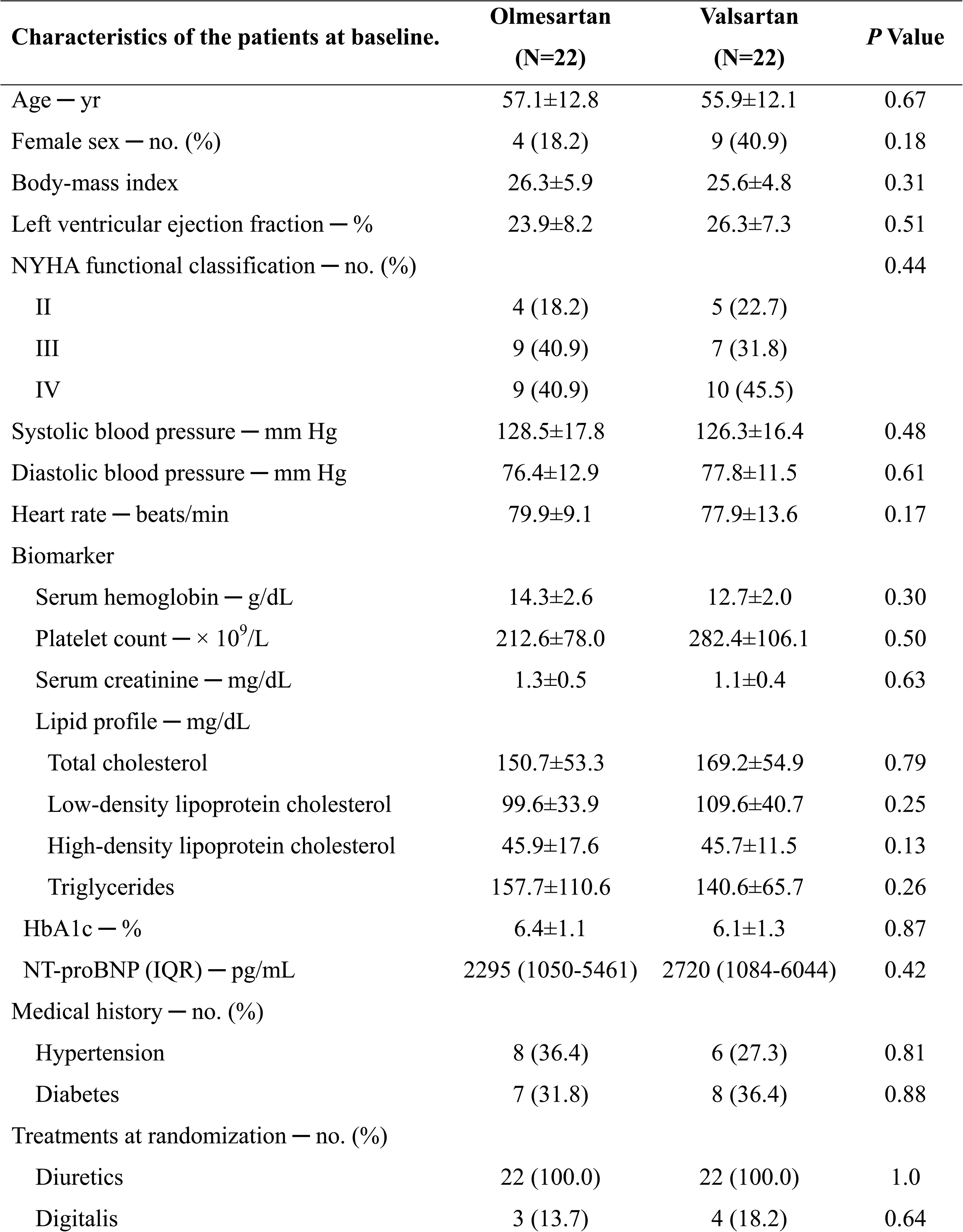
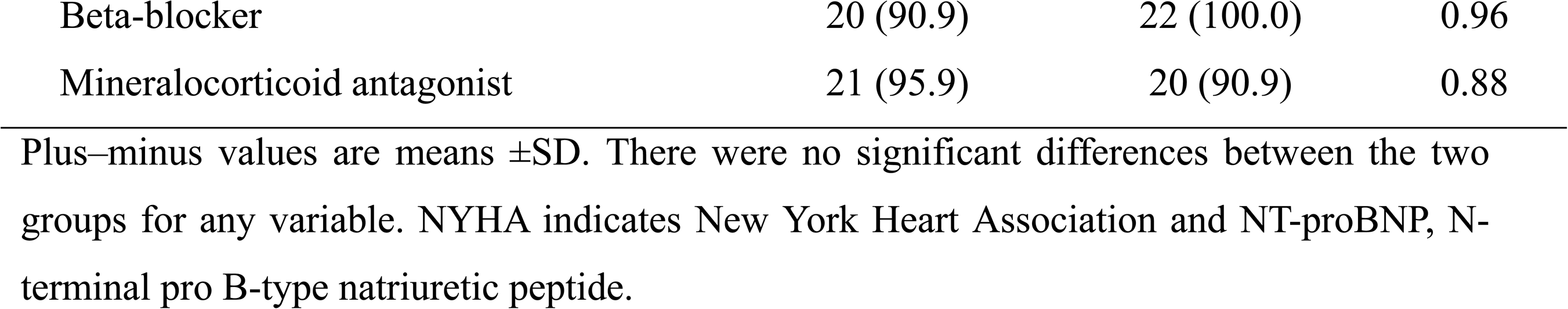

| Characteristics of the patients at baseline. | Olmesartan<br>(N=22) | Valsartan<br>(N=22) | *P* Value |
| --- | --- | --- | --- |
| Age — yr | 57.1±12.8 | 55.9±12.1 | 0.67 |
| Female sex — no. (%) | 4 (18.2) | 9 (40.9) | 0.18 |
| Body-mass index | 26.3±5.9 | 25.6±4.8 | 0.31 |
| Left ventricular ejection fraction — % | 23.9±8.2 | 26.3±7.3 | 0.51 |
| NYHA functional classification — no. (%) |  |  | 0.44 |
| II | 4 (18.2) | 5 (22.7) |  |
| III | 9 (40.9) | 7 (31.8) |  |
| IV | 9 (40.9) | 10 (45.5) |  |
| Systolic blood pressure — mm Hg | 128.5±17.8 | 126.3±16.4 | 0.48 |
| Diastolic blood pressure — mm Hg | 76.4±12.9 | 77.8±11.5 | 0.61 |
| Heart rate — beats/min | 79.9±9.1 | 77.9±13.6 | 0.17 |
| Biomarker |  |  |  |
| Serum hemoglobin — g/dL | 14.3±2.6 | 12.7±2.0 | 0.30 |
| Platelet count — × 10<sup>9</sup>/L | 212.6±78.0 | 282.4±106.1 | 0.50 |
| Serum creatinine — mg/dL | 1.3±0.5 | 1.1±0.4 | 0.63 |
| Lipid profile — mg/dL |  |  |  |
| Total cholesterol | 150.7±53.3 | 169.2±54.9 | 0.79 |
| Low-density lipoprotein cholesterol | 99.6±33.9 | 109.6±40.7 | 0.25 |
| High-density lipoprotein cholesterol | 45.9±17.6 | 45.7±11.5 | 0.13 |
| Triglycerides | 157.7±110.6 | 140.6±65.7 | 0.26 |
| HbA1c — % | 6.4±1.1 | 6.1±1.3 | 0.87 |
| NT-proBNP (IQR) — pg/mL | 2295 (1050-5461) | 2720 (1084-6044) | 0.42 |
| Medical history — no. (%) |  |  |  |
| Hypertension | 8 (36.4) | 6 (27.3) | 0.81 |
| Diabetes | 7 (31.8) | 8 (36.4) | 0.88 |
| Treatments at randomization — no. (%) |  |  |  |
| Diuretics | 22 (100.0) | 22 (100.0) | 1.0 |
| Digitalis | 3 (13.7) | 4 (18.2) | 0.64 |
| Beta-blocker | 20 (90.9) | 22 (100.0) | 0.96 |
| Mineralocorticoid antagonist | 21 (95.9) | 20 (90.9) | 0.88 |

Plus-minus values are means ± SD. There were no significant differences between the two groups for any variable. NYHA indicates New York Heart Association and NT-proBNP, N-terminal pro B-type natriuretic peptide.

### Study outcomes

^18^F-FDG PET data were successfully obtained for all patients at baseline. At baseline, SUVR was similar between the two groups (Table 2). Six-month follow-up ^18^F-FDG PET data were available in 40 (90.9%) patients. There was a significant between-group difference in the 6-month SUVR as the primary outcome. At the 6-month follow-up evaluation, SUVR decreased (*P* = 0.02) from 5.37 ± 3.33 to 3.65 ± 2.00 in the olmesartan group, but there was no difference (*P* = 0.56) in the valsartan group from 5.49 ± 3.65 to 5.51±2.50 (Figure 2). The magnitude of SUVR change was significantly different between the two groups (*P* = 0.02).

**Figure 2.**
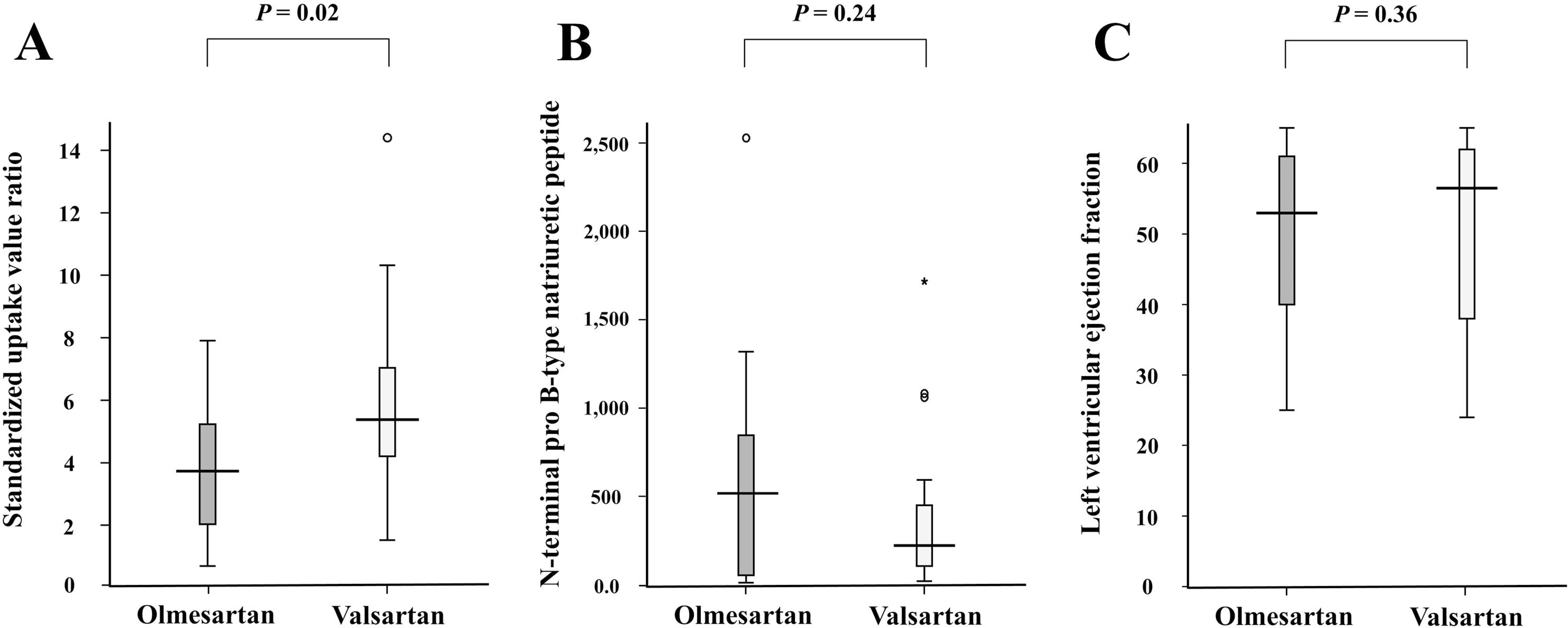
Changes in SUVR from baseline to 6 months of study medications. Plot illustrates the individual SUVR at baseline and after 6 months. SUVR indicates standardized uptake value ratio.

**Figure 3.**
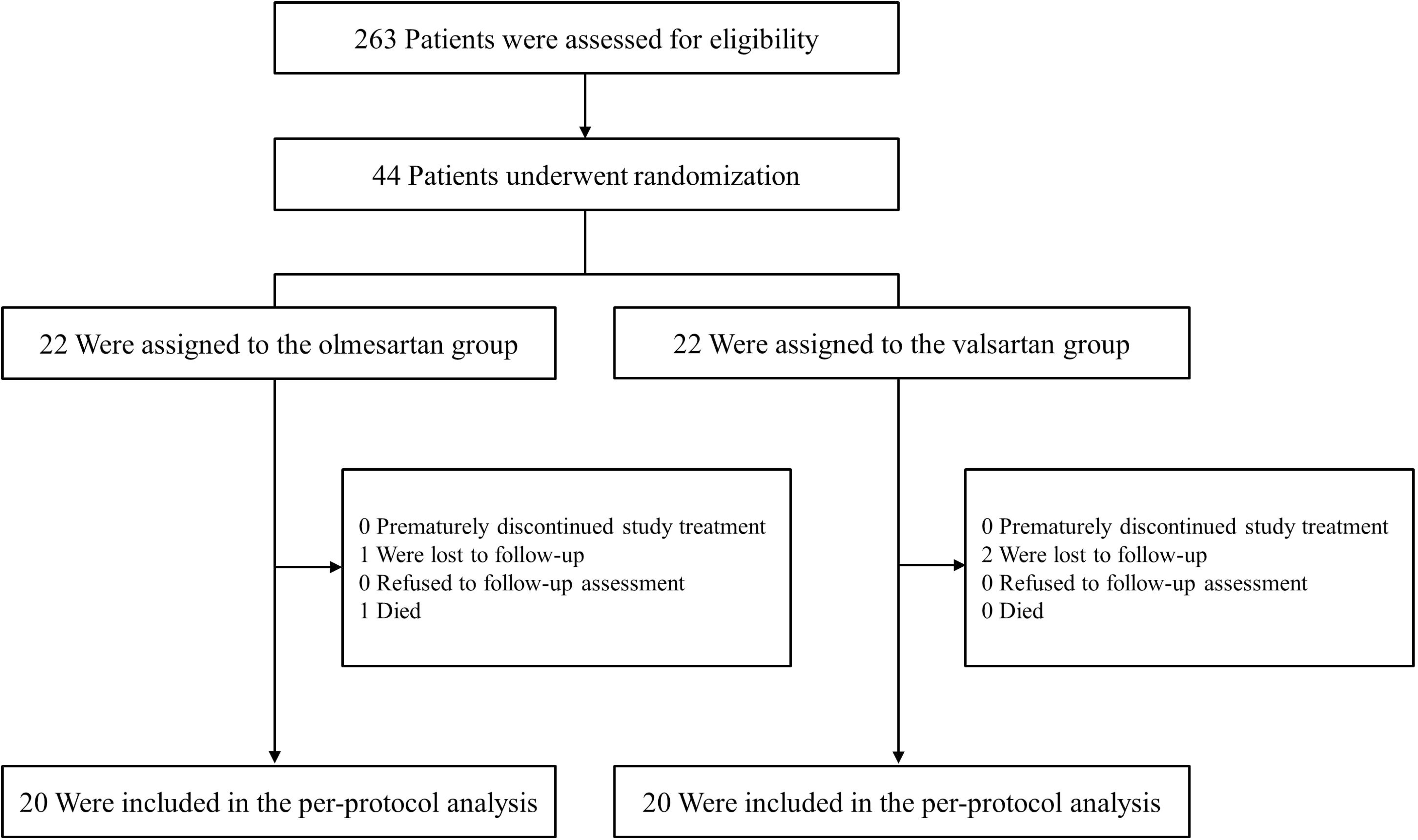
Effects of olmesartan and valsartan on the study end points. Panel A shows the SUVR between the study groups at 6 months, and Panel B shows the NT-proBNP. Panel C shows the LVEF over 6 months. The horizontal line within each box represents the median; the lower and upper borders of each box represent the 25^th^ and 75^th^ percentiles, respectively; and the T bars represent the differences between the lower and upper borders multiplied by 1.5. SUVR indicates standardized uptake value ratio; NT-proBNP, N-terminal pro B-type natriuretic peptide; and LVEF, left ventricular ejection fraction.

**Table 2.** Nuclear imaging between the olmesartan and valsartan groups.

| **Characteristic** | **Olmesartan** | **Valsartan** | ***P* Value** |
| --- | --- | --- | --- |
| Myocardial metabolism at baseline | **(N=22)** | **(N=22)** |  |
| Global SUVR | 5.37±3.33 | 5.49±3.65 | 0.91 |
| LAD territory SUVR | 5.25±3.34 | 5.40±3.66 | 0.89 |
| LCX territory SUVR | 5.54±3.35 | 5.73±3.72 | 0.86 |
| RCA territory SUVR | 5.31±3.32 | 5.49±3.65 | 0.86 |
| Myocardial metabolism at 6-month | **(N=20)** | **(N=20)** |  |
| Global SUVR | 3.65±2.00 | 5.76±3.10 | 0.02 |
| LAD territory SUVR | 3.53±1.97 | 5.58±2.89 | 0.01 |
| LCX territory SUVR | 3.91±2.23 | 5.91±3.22 | 0.03 |
| RCA territory SUVR | 3.52±1.88 | 5.81±3.21 | 0.01 |
| Change in SUVR | - 1.71±4.33 | 0.28±2.10 | 0.02 |

Plus-minus values are means ±SD. There were no significant differences between the two groups for any variable. SUVR indicates standardized uptake value ratio; LAD, left anterior descending artery; LCX, left circumflex artery; and RCA, right coronary artery.

For the secondary outcome, there was no significant between-group difference except for SUVR change. At 6 months, the level of NT-proBNP decreased significantly from baseline in both groups (both *P* < 0.01). However, the magnitude of NT-proBNP change was not significantly different between the two groups, and there was no difference in NT-proBNP levels (*P* = 0.24). At 6 months, the olmesartan group and the valsartan group showed a significant increase in echocardiography (50.0 ± 12.7 and 50.8±14.3, respectively) from baseline (both *P* < 0.01), although there was no significant difference between the two groups (*P* = 0.36). Clinical follow-up was completed for all patients in both groups at six months. One patient in the olmesartan group died of cardiac causes during the study period. No patient had adjudicated HF events, including hospitalization for HF or urgent HF visits, during the study period. No statistically significant associations between SUVR and other independent variables were observed (Table 3). Correlation analyses did not identify a significant relationship between SUVR and LVEF (*R* = - 0.25, *P* = 0.18) or NT-proBNP levels (*R* = - 0.15, *P* = 0.43).

**Table 3.** Univariable and multivariable regression analysis to determine factors associated with SUVR.

| Variable | Univariable |  | Multivariable |  |
| --- | --- | --- | --- | --- |
|  | Coefficient estimate | *P* Value | Coefficient estimate | *P* Value |
| Age | 0.01 | 0.48 | 0.03 | 0.60 |
| Female sex | - 0.34 | 0.63 | — | — |
| Body-mass index | 0.30 | 0.29 | 0.41 | 0.44 |
| Left ventricular ejection fraction | - 0.02 | 0.10 | - 0.04 | 0.30 |
| NYHA functional classification | 0.15 | 0.15 | 0.32 | 0.49 |
| Systolic blood pressure | 0.01 | 0.88 | — | — |
| Diastolic blood pressure | 0.02 | 0.70 | — | — |
| Heart rate | - 0.14 | 0.12 | - 0.29 | 0.21 |
| Serum hemoglobin | - 0.16 | 0.19 | - 0.35 | 0.32 |
| Serum creatinine | 0.67 | 0.57 | — | — |
| HbA1c | 0.36 | 0.22 | 0.46 | 0.51 |
| NT-proBNP | 0.01 | 0.39 | 0.03 | 0.68 |
| Hypertension | 0.28 | 0.72 | — | — |
| Diabetes | 0.22 | 0.54 | — | — |

SUVR indicates standardized uptake value ratio; NYHA, New York Heart Association, and NT-proBNP, N-terminal pro B-type natriuretic peptide.

### Safety outcomes

The incidence of adverse events in the olmesartan group was similar to that in the valsartan group. After randomization, two patients in the valsartan group had symptomatic hypotension. However, it did not require discontinuation of treatment during the study period.

## Discussion

The current study was the first to evaluate effects of ARB on myocardial glucose metabolism in patients with DCMP. First, we demonstrated that 6-month treatment with olmesartan decreased myocardial glucose metabolism, as measured by SUVR, by 48% compared to 6-month treatment with valsartan in patients with DCMP. Second, although SUVR was lower in olmesartan than valsartan, clinical outcomes were similar between these two ARBs over 6 months of treatment. Third, there were no significant differences in hemodynamic response between olmesartan and valsartan.

Myocardial metabolism is an integral part of the function of the heart as consumer and provider of energy. It has pleiotropic roles.^1,2^ Impaired cardiac metabolism and energy depletion are well recognized to contribute to cardiac dysfunction and reduced efficiency in terms of energy transfer to contractile work.^3–7^ These changes, which contribute to many cardiac pathologies by impairing ATP-dependent intracellular processes such as myofilament contraction and maintenance of ion homeostasis, have been described in prevalent disorders such as myocardial ischemia reperfusion injury, diabetes-related cardiac dysfunction, cardiac hypertrophy, and HF.^25^ Patients with DCMP exhibit myocardial metabolism dysfunction characterized by decreased fatty acid metabolism and increased myocardial glucose metabolism.^6,21^ However, this is less efficient and does not produce as much ATP. Those cardiac energy metabolic changes can manifest as both a deficit in energy production by the heart as well as a decrease in cardiac efficiency.^26,27^ Metabolic derangement in HF is not limited to the myocardium, but extends to skeletal muscles and contributes to the deterioration of exercise capacity in these patients, who can experience muscle weakness, fatigue, exercise limitation, dyspnea, and disease progression.^28^ Therefore, metabolic therapy can be an important mainstay for treating HF. Optimizing myocardial energy metabolism has been studied as a novel form of therapy.^29^ In early experimental studies, angiotensin II was involved in decreasing cardiac energy supply and cardiac efficiency.^8,9^ As a result, inhibition of RAAS might lead to improvement in myocardial metabolism.^10,11^ In this study, SUVR differences between olmesartan and valsartan might be caused by their different pharmacological properties. The antihypertensive action and duration of olmesartan might be greater than those of other ARBs because it is a more potent and selective angiotensin II receptor antagonist with no agonist activity.^30–33^ Olmesartan can block angiotensin II receptors more efficiently because angiotensin II does not increase. It can also enhance the effect of bradykinin through inhibition of kininase II. It has been reported that olmesartan can increase angiotensin converting-enzyme 2 expression in a remodeling heart after myocardial infarction, which theoretically could contribute to beneficial effects of ARB by facilitating increased cardiac Ang-(1-7) formation.^34^ The increase of Ang-(1-7) might have vasodilatory and organ-protective effects, such as inhibition of vascular remodeling and cardiac hypertrophy.^35–37^ Results of the present study suggest that each ARB might have different effects on myocardial targets that are very important for myocardial activity. However, relatively different contributions of myocardial metabolic effects of ARB to clinical benefits remain to be elucidated. The authors believe that the reason that the difference in SUVR did not appear as a clinical difference was probably due to the small number of registered patients.

Overactivity of the RAAS is associated with poor patient prognosis in DCMP.^38,39^ Previous studies have shown that chronic angiotensin II receptor stimulation can induce cardiac systolic dysfunction and electrical remodeling in the absence of hypertension.^40,41^ Angiotensin blockade is a major DCMP therapeutic strategy. It provides a balanced reduction in preload and afterload and inhibits cardiac hypertrophy and remodeling, therefore reducing mortality and morbidity.^16,42^ Valsartan can effectively inhibit the RAAS system. It is recommended for treating DCMP.^16,17,42^ Previous animal studies have demonstrated that olmesartan is effective in treating DCMP.^43,44^ The present study showed that olmesartan was as effective as valsartan in treating HF and improving LVEF and symptoms of DCMP.

It is important to consider limitations of our study. First, results obtained were derived from a small sample size with limited data not adequate enough to reflect clinical differences between olmesartan and valsartan in DCMP. In addition, due to the small sample size, study results might have been influenced by underlying physiological and pathological states of enrolled patients in the two study groups. Obesity, aging, and diabetes can cause heart metabolism alterations.^45–47^ Further study on whether the SUVR reduction effect by suppressing the RAAS is direct or indirect through some other factors is needed. Second, the heterogenicity of DCMP could not be ruled out as a confounding factor in study results. In this trial, additional diagnostic tests such as endomyocardial biopsy, bioptates assessment, and genetic studies were not performed. To minimize the heterogenicity of DCMP, we performed CAG to rule out coronary artery disease and excluded patients with primary valve disease or atrial fibrillation. Third, myocardial metabolisms of enrolled patients were evaluated using only ^18^F-FDG PET. Although ^18^F-FDG PET is currently the only evaluation tool for myocardial glucose metabolism, it is limited in evaluating differences in myocardial metabolism through changes in glucose metabolism among various types of metabolic changes.

In conclusion, six months of olmesartan therapy significantly decreased myocardial glucose metabolism based on measured SUVR in DCMP patients compared to six months of valsartan therapy. Further study with a longer duration and a larger number of patients is needed to confirm different effects of valsartan and olmesartan on myocardial metabolism.

## Data Availability

Yes. We include a data availability statment in the manuscript.

## Acknowledgements

The authors thank Young Jin Jeong (Department of Nuclear Medicine, Dong-A University Hospital, Dong-A University College of Medicine) for his advice on ^18^F-FDG PET data analysis.

## Funding Sources

This study was funded by a clinical research grant from DaeWoong Pharmaceutical and Dong-A University Research fund.

## Conflict of Interest Disclosures

None.

## List of abbreviations

ARB: Angiotensin II receptor blocker
CAG: coronary angiography
DCMP: Dilated cardiomyopathy
FDG: fluoro-2-deoxyglucose
HF: Heart failure
IQR: interquartile range
LVEF: Left ventricular ejection fraction
NT-proBNP: N-terminal pro-B-type natriuretic peptide
NYHA: New York Heart Association
OVOID: A comparison study of Olmesartan and Valsartan On myocardial metabolism In patients with Dilated cardiomyopathy
PET: Positron emission tomography
RAAS: Renin-angiotensin-aldosterone systems
SUV: standardized uptake value
SUVR: standardized uptake value ratio
VOI: Volume of interest

## Notes

### Competing Interest Statement

The authors have declared no competing interest.

### Clinical Trial

ClinicalTrials.gov; NCT04174456; 18 November 2019

### Funding Statement

We include details of all funding supported in the work presented.

### Author Declarations

The Ethics Committee/Institutional Review Board of Dong-A University Hospital.

